# The impact of physician handoffs on the outcomes of admitted emergency department patients: a medical administrative database retrospective cohort study

**DOI:** 10.1101/2025.01.16.25320692

**Authors:** Akram Mokhtari, David Simonyan, Myriam Mallet, S. Blais, L. Moore, Simon Berthelot

**Affiliations:** Université de Montréal, Montréal, QC, CAN; Centre de recherche du CHU de Québec-Université Laval, Québec, QC, CAN; CHU de Québec-Université Laval, Québec, QC, CAN

**Keywords:** Emergency department, handoff, mortality, cohort study

## Abstract

**Introduction:** A physician handoff is the process through which physicians transfer the primary responsibility of a care unit. The emergency department (ED) is a fast-paced and crowded environment where the risk of information loss between shifts is significant. Yet, the impact of handoffs between emergency physicians on patient outcomes remains understudied. We performed a retrospective cohort study in the ED to determine if handed-off patients, when compared to non-handed-off patients, were at higher risk of negative outcomes.

**Methods:** We included every adult patient first assessed by an emergency physician and subsequently admitted to hospital in one of the five sites of the CHU de Québec-Université Laval during fiscal year 2016-17. Primary outcome was mortality. Secondary outcomes were incidence of ICU admission and surgery and hospital length of stay. We conducted propensity score based analysis accounting for patient and hospital clusters and adjusting for demographics, multiple disease severity indicators and ED processes indicators, including crowding.

**Results:** 21,136 ED visits and 17,150 unique individuals were included in the study. Median[Q1-Q3] age, Charlson score, door-to-emergency-physician time and ED length of stay were 71[55-83] years old, 3[1-4], 48 [24,90] minutes, 20.8[9.9,32.7] hours, respectively. In propensity score analysis (OR handoff/no handoff [CI95%] or GMR[Cl95%]), handoff status was not associated with mortality (1.08[0.93,1.26]), ICU admission (1.01[0.87,1.18]) or hospital length of stay (1.02[0.94-1.10]). Sensitivity and sub-group based analyses yielded no further information.

**Conclusion:** Emergency physicians’ handoffs were not associated with an increase in risk of severe in-hospital adverse events. Further studies are needed to explore the impact of ED handoffs on adverse events of low and moderate severity.

**What is already known on this subject:** Handoffs are widely believed to affect patient outcomes in the ED, although data remains scarce.

**What did this study ask:** What is the impact of handoffs between emergency physicians on patient outcomes

**What this study adds:** Handed off patients do not seem to do worse than non handed off patients concerning majors outcomes (mortality, ICU admission, LOS).

**Why does it matter to clinicians?:** In the ED, handoffs may not represent a risk to patient outcomes. QI efforts may be better invested in other care inefficiencies.

## INTRODUCTION

A physician handoff is a transfer and acceptance of patient care responsibility between a physician finishing a clinical duty and another physician taking over that clinical duty(1). Some data seem to indicate that handoffs may pose a risk to patient care through a break down in communication (2).

This potential risk may especially be problematic in fields like emergency medicine, which operate fully under shift work. The risk of a suboptimal transmission or a loss of information in the emergency department (ED) is high; the environment is stressful; the caseload is often heavy and patient care is frequently shared among multiple physicians. This type of care environment may lead to a suboptimal transfer of critical information on patients at higher risk of poor health outcomes (3).

Some studies have evaluated ED handoffs. Polls and observational studies seem to indicate that ED physicians generally believe handoff are risk to patient outcomes (4-6) and that potentially key information is omitted in up to 30-45% of handoffs (7-9) Previous studies have shown an association between other suboptimal care processes (i.e crowding and time to care) and unfavorable outcomes, including mortality(10-12). However, data on the impact of inadequate handoffs on direct patient outcomes are lacking(13).

To address this major knowledge gap, this study aimed to compare the outcomes of patients that have been involved in a patient handoff in the emergency department to those of patients with no handoff.

## METHODS

### Study Design

We conducted a retrospective, multicenter cohort study. This project was approved by the Ethics committee of the CHU de Québec – Université Laval.

### Setting and Population

This study is set in the healthcare system of the province of Quebec, Canada, a fully mature public healthcare system. The study includes the five EDs and hospitals that form the CHU de Québec-Université Laval (CHUQ-UL), one of the three largest academic health institutions in Canada. The five ED sites of the CHUQ-UL receive more than 230,000 visits per year and serve the population of Québec city, eastern Québec and northeastern New Brunswick, a catchment area of nearly 2 million people(14). The CHUQ-UL provides comprehensive care with specialties ranging from primary to quaternary care.

The study population includes adult patients (>18 years old) that were hospitalized through any of the emergency departments of the CHUQ-UL between April 2016 and March 2017. To be eligible, the patient’s first physician-led evaluation in the emergency department had to be conducted by an emergency physician. Patients initially assessed in other institutions and secondarily transferred to one of the participating sites were excluded

### Data sources

Data were extracted and combined by the clinical and organisational performance management unit of the CHUQ-UL, a unit regularly involved in performance studies at local, regional and national levels.

Data were extracted from two clinical and administrative databases available in Québec: MED-ECHO and Siurge. The MED-ECHO database holds data captured from all hospitalizations in the province. We extracted information on patient characteristics (e.g. age, sex) in-hospital care and diagnoses at hospital discharge. From the Siurge database, we extracted detailed information regarding ED care including ED operations (e.g. occupation rate, timing of clinical and para-clinical tests, admission requests), care provided and diagnoses at ED discharge. Data from both databases were matched using provincial unique patient codes (RAMQ).

### Outcomes

Our primary outcome was in-hospital mortality. Secondary outcomes were admission to the intensive care unit, surgery and hospital length of stay. Hospital length of stay was defined as the period between admission from the emergency department and hospital discharge. These measures were selected to align our study with previous literature evaluating the impact of ED operations on patient outcomes. (15, 16).

### Definition of Handoff (Figure 1 and Appendix 1)

In our study, patients were classified as either handed off or not handed off. Handoff status was determined based on the clinical patterns within the CHUQ-UL and clinical expertise. The four factors taken into account were 1) definition of episode of care 2) shift-work structure and allocation 3) handoff method 4) patient care responsibility. Within the CHUQ-UL, shifts last 8 hours and shift change happens at 8:00, 16:00 and 24:00. An episode of care was defined as the time between evaluation by an emergency physician and evaluation by the admitting specialist, as this the period in which the patient is under direct responsibility of an emergency physician. Handoffs are conducted orally, on an informal basis. No standardized handoff method, such as checklists, IT, security checks, are used within the study hospital system (Appendix 1). Also, within the CHUQ-UL, the leaving physician has no obligation to complete his caseload before transferring the responsibility of the emergency room. Lastly, we took into account that patients seen by an admitting specialist within the first hour of a shift are very rarely reevaluated by an emergency physician within that time frame.

**Figure 1.**
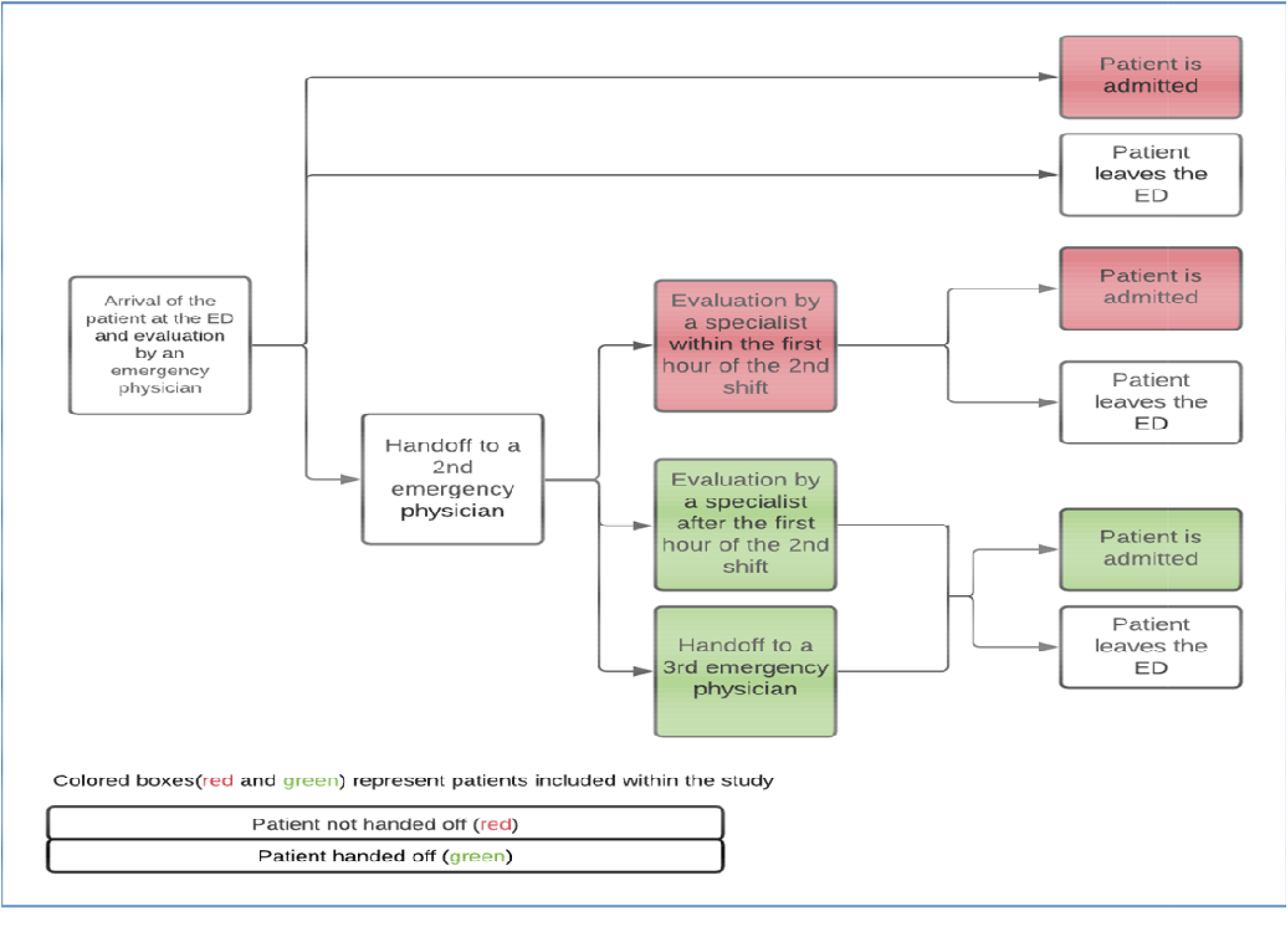
Flowchart of the possible clinical pathways for a patient within the study.

A patient was considered handed off if:

1. his episode of care period overlapped partially or totally with at least two emergency physician shifts
2. the time of his evaluation by an admitting specialist was, at the earliest, one hour after the beginning of the second shift in which the patient was in the ED

In all other possible clinical pathways, the patient was classified as not handed off.

### Statistical analysis (Appendix 2)

Mortality, admission to the intensive care unit and surgery incidence were compared across the two study groups using odd ratios (OR) and 95% confidence intervals [CI] generated with logistic regression models. Hospital length of stay was compared using geometric mean ratios (GMR) and 95% CI generated using log-linear models. We used random intercept multi-level regression models to account for patient-based and hospital-based clusters. In primary analysis, we used a propensity score to adjust OR/GMR for patient risk factors. We estimated the propensity of handoff using logistic regression based on covariates that were identified and selected *a priori* using literature review and clinical expertise. We thus adjusted for the following patient risk factors: age, sex, comorbidities (Charlson Index)(17) and diagnoses on hospital discharge. Regarding care processes, we adjusted for initial ED orientation (resuscitation, stretcher, ambulatory), Canadian Triage and Acuity Scale (CTAS) score, admitting department (medicine, surgery, family medicine), delay before evaluation by an emergency physician, delay before evaluation by another speciality, emergency department crowding and handoff timing (day, evening, night). Delay before evaluation by a specialist was defined as the delay between the first evaluation by an emergency physician and the first evaluation by an admitting specialist.

ED crowding was estimated using two parameters: 1) average number of new ED visits per hour during the ED care episode of each patient; and 2) average ED occupancy rate (in patient presence time), divided by the number of allowed beds for each emergency department. Patient presence time is, for any given hour, the total amount of time patients have spent in the emergency department. For example, two patients staying for thirty minutes would account for a total of 1 patient-hour.

In supplemental analysis, we conducted regular logistic/log-linear regression analysis to assess the specific effect of each confounder on different outcomes.

### Sensitivity analyses

To explore the validity of our handoff status definition, we conducted a sensitivity analysis in which we defined handoff status solely based on the official shift cut-offs (8:00, 16:00 and 24:00). In this analysis, a patient was considered handed off if his ED episode of care overlapped with at least two different emergency physician shifts.

We conducted the following additional sensitivity analyses:

- Excluding patients who had the outcome (death, ICU admission,) i) > 72 hours or ii) > 7 days after admission to account for a potential difference in time of occurrence of events between study populations.
- Excluding patients who had an ED length of stay > 24h to account for the possibility that an extended ED stay is due to external factors (e.g awaiting placement in a long-term care facility)

### Subgroup analyses

We conducted subgroup analyses to explore potential variations based on aging (<50, 50-64, 65-74, 75-84, 85+) and hospital site care patterns (analysis by hospital)

## RESULTS

### Patient population (Table 1)

The study population included all 21,136 emergency room visits that resulted in hospital admission. There were 17,150 unique individuals, for a hospital admission proportion of 13%. A patient handoff occurred in 49.4% of emergency room visits. Forty-two percent of patients were older than 75 years old and 60.1% of patients had a Charlson index score of three or more. Median (IQR) delay before evaluation by an emergency physician was 48 (24;90) minutes.

**Table 1.** Characteristics of study patients by handoff status.

| N(%) or<br>Median (Q1;Q3) | *Not handed off*<br>n=10,443 | *Handed off*<br>n=10,693 | *Total*<br>n=21,136<sup>#</sup> |
| --- | --- | --- | --- |
| **Sex** |  |  |  |
| Male | 4,243 (49)* | 3,695 (43)* | 7,938 (46)* |
| *analysis by unique patient (N=17148) |  |  |  |
| **Age** |  |  |  |
| 18-49 | 1,939 (23)* | 1,376 (16)* | 3,315 (19)* |
| 50-64 | 1,770 (21)* | 1,477 (17)* | 3,247 (19)* |
| 65-74 | 1,710 (20)* | 1,666 (19)* | 3,376 (20)* |
| 75-84 | 1,678 (20)* | 2,050 (24)* | 3,728 (22)* |
| 85+ | 1,461 (17)* | 2,021 (24)* | 3,482 (20)* |
| *analysis by unique patient (N=17148) |  |  |  |
| **Charlson Index** |  |  |  |
| 0 | 1,749 (20)* | 1,220 (14)* | 2,969 (17)* |
| 1 | 943 (11)* | 745 (9)* | 1,688 (10)* |
| 2 | 1,169 (14)* | 1,000 (12)* | 2,169 (13)* |
| 3 | 1,380 (16)* | 1,455 (17)* | 2,835 (17)* |
| 4 | 1,865 (22)* | 2,160 (25)* | 4,025 (23)* |
| 5 | 935 (11)* | 1,347 (16)* | 2,282 (13)* |
| 6+ | 517 (6)* | 652 (7)* | 1,169 (7)* |
| *analysis by unique patient (N=17148) |  |  |  |
| **Emergency department**<br>**length of stay (hours)** | 11.5 (6.5;26.4) | 25.5 (17.8;42.4) | 20.8 (9.9;32.7) |
| **Delay before evaluation**<br>**by an emergency physician (hours)** | 0.7 (0.3;1.3) | 1.0 (1.0;1.0) | 0.8 (0.4;1.5) |
| **Delay before evaluation by a**<br>**specialist (hours)** | 2.4 (1.3;3.9) | 12.6 (8.1;17.3) | 5.2 (2.3;12.8) |
| **Canadian Triage and Acuity Scale** |  |  |  |
| 1 | 1054 (10) | 335 (3) | 1389 (7) |
| 2 | 2392 (23) | 1933 (18) | 4325 (20) |
| 3 | 4417 (42) | 5215 (49) | 9632 (46) |
| 4 | 2185 (21) | 2991(28) | 5176 (25) |
| 5 | 395 (4) | 219 (2) | 614 (3) |
| **Initial orientation in the ED** |  |  |  |
| Ambulatory | 56 (0.5) | 8 (0.1) | 64 (0.3) |
| Non resuscitation stretcher | 7991(76.5) | 9739 (91) | 17730 (83.9) |
| Resuscitation bay | 2396 (23.0) | 946 (8.9) | 3342 (15.8) |
| **Admitting specialty category** |  |  |  |
| Medicine | 7026 (67) | 6997 (65) | 14023 (66) |
| Surgery | 3099 (30) | 2505 (23) | 5604 (27) |
| Family medicine | 318 (3) | 1191 (11) | 1509 (7) |
| **Time of handoff** |  |  |  |
| Morning shift 8:00 | N/A | 2159 (20) | N/A |
| Evening shift 16:00 | N/A | 3214 (30) | N/A |
| Night shift 24:00 | N/A | 5320 (50) | N/A |
| **Number of handoffs** |  |  |  |
| 0 | N/A | 10443 | N/A |
| 1 or 2 | N/A | 7328 | N/A |
| 3 | N/A | 2514 | N/A |
| 4 or more | N/A | 851 | N/A |
| **Average number of new ED visits during episode of care per hour per allowed bed** | 0.22 (0.16;0.26) | 0.17 (0.13;0.22) | 0.19 (0.14;0.24) |
| **Average ED occupancy rate during episode of care per allowed bed (patient-hour)** | 1.59 (1.40;1.79) | 1.51 (1.30;1.71) | 1.55 (1.34;1.75) |

Median (IQR) ED length of stay varied across hospital sites; the longest being 33.8 (32.9; 34.6) hours and the shortest being 19.8 (19.4; 20.2) hours.

Handed off patients tended to be older, have more comorbidities (Charlson index ≥ 2; 78% vs. 69%) and lower disease severity (CTAS score 1-2; 21% vs 33%). Further, handoffs were more frequent in patients presenting during the evening shift (16:00-24:00) while non-handed off patients tended to visit during the day shift (8:00-16:00). Handed off patients had a significantly longer median ED length of stay (25.5h vs 11.5h).

### Mortality (Tables 2-3)

In our cohort within the study period, 1,819 in-hospital deaths (8.6%) were reported. Handoff status was not associated with incidence of death in crude or propensity score analysis.

**Table 2.** Descriptive outcome data by handoff status.

| N(%) or<br>Median (Q1;Q3) | *Not handed off*<br>**n=10,443** | *Handed off*<br>**n=10693** | *Total*<br>**n=21136** |
| --- | --- | --- | --- |
| **In-hospital mortality** | 981 (9.2) | 837 (7.8) | 1818 (8.6) |
| **Admission to the ICU** | 1407 (13.4) | 642 (6.0) | 2049 (9.7) |
| **Hospital length of stay** | 6 (3.0 - 13.0) | 6 (3.0 - 15.5) | 6 (3 - 14) |

**Table 3.** Odds ratios (OR)* and geometric mean ratios (GMR)* for bivariate and propensity score analyses by handoff status.

|  | *Mortality(OR)*<br>(OR [95% CI]) | *ICU admission(OR)*<br>(OR [95% CI]) | *Length of stay(GMR)*<br>(OR [95% CI]) |
| --- | --- | --- | --- |
| **Bivariate\*\*** | 0.77 (0.70-0.85) | 0.38 (0.35-0.43) | 1.18 (1.13-1.23) |
| **Propensity score** | 1.08 (0.93-1.26) | 1.01 (0.87-1.18) | 1.02 (0.94-1.10) |

\*OR or GMR > 1 imply higher odds for handed off patients; <1 imply lower odds for handed off patients
\*\*Crude analysis taking into account hospital and patient clusters

#### Sensitivity and sub-group analyses (Table 4)

Age and hospital sub-group analyses did not lead to significantly different results. Hospital and ED length of stay sensitivity analyses yielded no further information. Subgroup analyses by age and hospital yielded no further information.

**Table 4.**
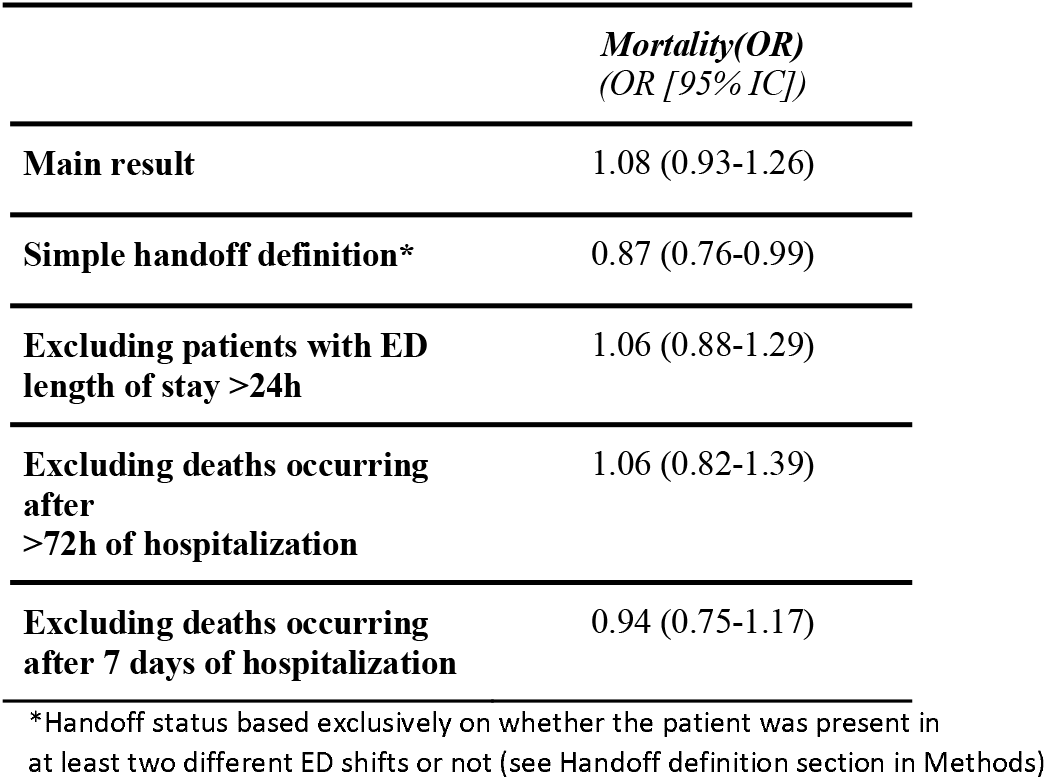
Sensitivity analyses of the main outcome.

|  | *Mortality(OR)*<br>(OR [95% IC]) |
| --- | --- |
| **Main result** | 1.08 (0.93-1.26) |
| **Simple handoff definition\*** | 0.87 (0.76-0.99) |
| **Excluding patients with ED length of stay >24h** | 1.06 (0.88-1.29) |
| **Excluding deaths occurring after >72h of hospitalization** | 1.06 (0.82-1.39) |
| **Excluding deaths occurring after 7 days of hospitalization** | 0.94 (0.75-1.17) |

\*Handoff status based exclusively on whether the patient was present in at least two different ED shifts or not (see Handoff definition section in Methods)

In sensitivity analysis, we compared our study handoff definition to a definition based exclusively on official shift cut-offs (8:00; 16:00; 24:00). In the latter, handed off patients had statistically significant lower odds of death than non-handed off patients (OR=0.87 [0.76,0.99]).

### Intensive care unit admission (Table 2-3)

Within study period, 2,049(9.7%) events were reported. In crude analysis, handed off patients had a statistically significantly lower odds of ICU admission (OR=0.38[0.35-0.43]). However, handoff status was not associated with ICU admission incidence in propensity score analysis.

### Hospital length of stay (Table 2-3)

Median hospital length of stay was of 6 (IQR) days for both handed off (3.0-15.5) and non-handed off patients (3.0-13.0). In propensity score analysis, handoff status was not associated with hospital length of stay.

### Associations between confounding variables and outcomes

Multivariate analyses demonstrated that, unlike being transferred from one emergency physician to another, older age, the presence of comorbidities, initial treatment in the ED resuscitation bay, the stretcher occupancy rate in the ED, and a shorter ED length of stay are all independently associated with higher mortality and an increased risk of admission to intensive care. The results of these additional analyses are available in Appendix 3.

## DISCUSSION

### Interpretation of findings

Handed off patients tended to be older, have a higher comorbidity burden, but lower acute disease severity. In propensity score analysis, handoff status was not associated with any of the study outcomes.

At baseline, non-handed off patients are at higher risk of short-term unfavorable outcomes. Indeed, these patients tend to be more acutely ill individuals that are rapidly transferred to higher levels of care and do not stay in the emergency department as long. This trend is confirmed by the results of the present study as, in univariate analysis, non-handed off patients had a higher risk of death and ICU admission. When taking into account multiple confounding factors including disease severity, both patients groups in our sample had a similar risk of adverse events.

### Comparison to previous studies

Previous retrospective studies reviewing malpractice claims, have linked handoffs to a variety of failures of care including intensive care unit complications or death(16, 18). This seems to contradict our findings. Firstly, these types of studies do not weight the contribution of the handoff compared to the multitude of other factors that are part of the chain of events of an adverse outcome. Finally, on a case to case basis, a suboptimal handoff may lead to an adverse event, but, these situations may not be frequent enough to produce a difference at a population level.

### Study handoff definition

The handoff definition used in this study was based on clinical expertise and local clinical patterns to ensure our study reflected real emergency medical practice. However, we believe the definition employed, considering any patient that was evaluated by an admitting specialist within the first hour of a shift as not handed off, reflects the general reality of most ED across North America and other modern healthcare systems. Indeed, at the beginning of a shift, the leaving emergency physician often stays to complete unfinished tasks and the entering physician has to prepare for his shift and see new patients(19, 20). Handoffs are thus very frequently delayed. Further, when an admitting specialist is about to arrive, the entering physician is unlikely to re-evaluate that patient as 1) the disposition of that patient has been more or less decided 2) high patient volume in most EDs delays patient re-evaluations(3). However, handoff methods are heterogeneous and our definition may not apply everywhere. We conducted a sensitivity analysis using exclusively the shift cut-offs to determine handoff status. In that analysis, handed-off patients had a marginally lower odds of mortality (0.87[0.76,0.99]), which is coherent with their higher acute risk profile.

### Strengths and limitations

Study data originated exclusively from administrative databases. The validity of our results are thus dependent on redaction and coding quality of patient data. Mistakes in information entry may have lead to an underestimation of the OR/GMR because of a non-differential information bias. However, previous studies have shown that local administrative databases provide highly reliable data(21, 22). In our study, only ED presentations that resulted in hospitalisation were included. This may limit the extension of our findings to the non-admitted emergency room population. Admitted patients tend to be sicker and have a more complex medical profile. These patients are therefore the most at risk of suffering from unfavorable outcomes secondary to a loss of information.

Regarding strengths, as we included every admitted patient during the study period, this study is population-based. Further, study power enabled us to take account of an extensive number of confounding factors in terms of patient and disease characteristics and care processes. In addition, propensity score adjustment helped further reduce potential residual confounding by accounting for the different factors that can influence exposure to a handoff (e.g. ED crowding). Finally, the studied outcomes were direct patient outcomes which permitted us to evaluate whether handoffs had a concrete impact on patients.

### Clinical and research implications

There is a general belief in the emergency medicine community that handoffs represent a serious risk to patient outcomes, even if evidence is lacking(13). Consequently, a large number of initiatives have been conducted to standardize handoffs, with neutral or even detrimental outcomes(23, 24).

Our study seems to indicate that handoffs may not represent a significant risk to patient care in emergency medicine. Thus, it provides the basis to conduct prospective studies to confirm the trend observed in our study and potentially reorient research and clinical efforts and resources towards more risky processes (i.e crowding).

## CONCLUSION

In this retrospective cohort study of admitted patients, we did not observe an association between handoffs and mortality, ICU admission or hospital LOS. Our results do not address low morbidity consequences or patient safety events. Current efforts to standardize hand offs may therefore be suboptimal. To confirm our observations, further prospective studies evaluating the impact of handoffs on high and low morbidity consequences (e.g. overdiagnosis, overtreatment, resource use) are needed.

## Data Availability

All data produced in the present study are available upon reasonable request to the authors

