## Appendix 1 for "The impact of physician handoffs on the outcomes of admitted emergency department patients: a medical administrative database retrospective cohort study"

**Appendix 1- Detailed explanation of handoff status definition**

| **Key Concept** | **Definition** | |
| --- | --- | --- |
| Study care episode | Period beginning at the first evaluation by an emergency physician and ending at the first evaluation by an admitting specialist. This period is the period of interest, because it is the period during which a patient is under the direct responsibility of an emergency physician and at risk of information handoff failure. | |
| Shift-work structure and allocation | Work shifts last 8 hours and shift changes happen at 8:00, 16:00 and 24:00. Further, 10 minutes before the end of a shift, the leaving physician stops seeing new patients. Within that period, any new patient is seen by the physicians of the next shift. Therefore, shifts effectively start and finish 10 minutes earlier than theoretically planned. | |
| Handoff method | The handoff method employed is the case transfer. The leaving physician^1^ has no obligation to complete his caseload before transferring the responsibility of the emergency room to the entering physician^2^. There are no standardized time or method to proceed with the transfer. Patient information and care responsibility are transferred within the first hour of the shift of the entering physician. | |
| *Patient care responsability* | The entering physician is responsible for all the patients which were transferred to him. However, when a patient is seen by an admitting specialist within the first hour of a shift, the emergency physician will not, except in rare circumstances, re-evaluate that patient for two reasons: A) These patients are *de facto* under the responsibility of the admitting specialist. B) Official handoff happened after the beginning of the shift which delays patient re-evaluation. Therefore, when a patient was seen by an admitting specialist within the first hour of a shift, that patient was not considered as handed off between two emergency physicians, as the entering physician was never involved in the care of this patient. | |
| Handoff | A patient was considered handed off if | - His episode of care period overlapped partially or totally with at least two emergency physician shifts |
|  |  | - The time of his evaluation by an admitting specialist was, at the earliest, one hour after the beginning of the second shift in which the patient was in the emergency room |
| 1.Physician that is ending a shift at emergency department. A leaving physician is the physician that will handoff responsibility of his patients to a colleague.  2.Physician that is beginning a shift at the emergency department. An entering physician is the physician that will receive responsibility of patients in a handoff. | | |
