## Appendix 2 for "The impact of physician handoffs on the outcomes of admitted emergency department patients: a medical administrative database retrospective cohort study"

**Appendix 2- Propensity score and variable definition**

Propensity score

Logit_Handoff =  β_1 age_ + β_2 sex_ + β_3_ _c-index_ + β_4 ED-orientation_ + β_5_ _CTAS_ + β_6_  _Admitting Dept_ + β_7 D_MD_URG_

+ β_8_ _D_MD_URGSPE_ + β_9 D_URG_ β_10_ _Occupancy_ + β_11_ _New visits_ + β_12_ _Timing_

*Legend*

Age = age of the individual

Sex = biological sex of the individual

c-index= Charlon Index

ED-orientation = Initial orientation of the patient within the ED as either ambulatory, stretcher or reanimation bay

CTAS= Canadian triage and acuity score

Admitting Dept = Specialty admitting the patient

D_MD_URG = Delay before evaluation by an emergency physician

D_MD_URGSPE= Delay between evaluation by an emergency physician and evaluation by an admitting specialist

D_URG= Total length of stay in the ED from triage to admission.

Occupancy= Average patient-presence time per allowed bed during the patient’s ED stay (see below)

New visits= Average number of new visits per hour per number of allowed beds during the patient’s ED stay (see below)

Timing= Time of handoff (morning handoff, PM handoff, night handoff) as defined by the cutoff between shifts (8:00, 16:00, 24:00)

**Variables Occupancy and New visits**

These two variables were used to estimate crowding in our study.

An important notion that was used to estimate crowding with these two variables was theoretical ED capacity (in number of beds). For a given ED, its theoretical ED capacity is the number of patients that would be allowed in that ED under normal circumstances (no crisis, no overcrowding). This capacity is determined by the local health authorities based on the number of beds and caregivers available and capacity to offer optimal care to each patient. If the number of patients in the ED exceeds that value, the ED is considered overcrowded or in overcapacity. For example, for a given ED with a theoretical capacity of 30 beds, if there are 40 patients receiving care in that ED, that ED is considered in overcapacity.

Occupancy was determined based on the average patient-presence time for every given hour during the length of stay of a patient divided by the number of allowed beds. For example, if between 1:00 pm and 2:00 pm, there were 5 patients in the ED, 4 of them staying the whole hour and 1 of them staying only between 1:00 and 1:30 pm, the total patient-presence time for that given hour was of (4 patients x 1 hours of presence + 1 patient x 30 mins of presence) 4:30 hours. That number was then divided by the theoretical maximum capacity of the ED. This number is determined by the regional health authorities. For example, for a given ED that has a maximal capacity of 3 beds, we divided by 3. Note that EDs are frequently in over-capacity and we made this calculus to account for that factor in our crowding assessment.

We then averaged the occupancy rate of each hour of the stay of the patient to obtain the average occupancy rate while the patient was present.

Complete example:

Emergency department A with a theoretical ED capacity of 30 beds. Between 1:00 pm and 2:00 pm there were 40 patients receiving care in the ED.

Number of patients in the ED: 40 (30 stayed the full hour while 10 stayed for 30 minutes)

Patient presence time: (30x1h + 10 x 0,5 hours) 35 patient-hours

Occupancy rate: 35 patients-hours divided by a capacity of 30 beds = 1.17 patients-hours/bed

New visits was used to account for the pressure of multiple new visits on care given in the ED. News visits was determined based on the number of visits per hour for every given hour of a patient’s ED stay divided by the theoretical bed capacity of that ED. If between 1:00 pm and 2:00, 10 new patients arrived in the ED, that accounted for 10 new visits. If the given capacity of that ED is of 3 beds, we then divided by 3 We then averaged the number of new visits of each hour of the stay of the patient to obtain the average number of news visits.

Complete example:

Emergency department A between 1:00 pm and 2:00 pm with theoretical ED capacity of 30 beds. Between 1:00 and 2:00 pm there were 10 new patient visits at the ED.

Number of new visits: 10

New visits= (10 new visits / 30 bed) = 0,3 new visits/be
