## Appendix 3 for "The impact of physician handoffs on the outcomes of admitted emergency department patients: a medical administrative database retrospective cohort study"

**Appendix 3- Logistic regression analysis**

**Adjusted Odds Ratios (OR) and geometric means ratio (GMR) in full model analysis (all variables included) for mortality, ICU admission, surgery and length of stay**

|  | ***Mortality(OR)***  *(beta [95% IC])* | ***ICU admission(OR)***  *(beta [95% IC])* | ***Surgery(OR)***  *(beta [95% IC])* | ***Length of stay(GMR)***  *(GMR [95% IC])* |
| --- | --- | --- | --- | --- |
| **Male** | 1.09 (0.98-1.21) | 1.18 (1.06-1.31) | 1.01 (0.92-1.09) | 0.94 (0.90-0.98) |
| **Age**  18-49  50-64  65-74  75-84  85+ | 0.77 (0.46-1.30)  1.00  1.46 (1.19-1.79)  1.69 (1.34-2.12)  2.51(1.98-3.18) | 1.35 (0.99-1.85)  1.00  0.80 (0.66-0.96)  0.52 (0.42-0.65)  0.30 (0.24-0.39) | 1.27 (0.95-1.71)  1.00  1.00 (0.84-1.19)  0.99 (0.80-1.22)  0.88 (0.70-1.11) | 1.01 (0.86-1.18)  1.00  1.16 (1.07-1.26)  1.33 (1.21-1.47)  1.47 (1.33-1.64) |
| **Charlson Index**  0  1  2  3  4  5  6 + | 1.00  2.86 (1.49-5.50)  5.85 (3.00-11.42)  5.12 (2.58-10.18)  6.83 (3.42-13.62)  7.10 (3.54-14.21)  8.92 (4.45-17.89) | 1.00  1.50 (1.09-2.07)  1.95 (1.38-2.78)  2.08 (1.43-3.04)  2.71 (1.84-4.01)  2.42 (1.60-3.66)  2.58 (1.70-3.94) | 1.00  1.09 (0.82-1.45)  0.94 (0.68-1.30)  0.94 (0.67-1.33)  1.07 (0.75-1.52)  0.93 (0.64-1.35)  0.81 (0.55-1.19) | 1.00  1.46 (1.25-1.71)  1.70 (1.44-2.02)  1.74 (1.46-2.07)  1.93(1.61-2.32)  2.33 (1.93-2.81)  2.36 (1.95-2.86) |
| **Emergency department length of stay** | 0.99 (0.98-0.99) | 0.95 (0.94-0.96) | 0.98 (0.96-0.98) | 1.01 (1.00-1.02) |
| **Delay before evaluation by an emergency physician (per hour)** | 0.94 (0.89-1.00) | 0.92 (0.85-0.99) | 0.98 (0.94-1.02) | 0.99 (0.97-1.01) |
| **Delay before evaluation by a specialist (per hour)** | 1.00 (0.99-1.01) | 1.00 (0.99-1.02) | 1.00 (0.98-1.01) | 1.00 (1.00-1.00) |
| **Canadian Triage and Acuity Scale**  1  2  3  4  5 | 1.97 (1.51-2.56)  0.94 (0.78-1.12)  0.88 (0.77-1.00)  1.00  0.78 (0.54-1.12) | 3.05 (2.37-3.91)  1.65 (1.36-2.00)  1.20 (1.02-1.41)  1.00  0.66 (0.42-1.03) | 0.59 (0.46-0.76)  0.75 (0.65-0.86)  0.93 (0.84-1.04)  1.00  1.18 (0.93-1.49) | 0.80 (0.71-0.91)  0.86 (0.80-0.92)  0.86 (0.81-0.90)  1.00  0.99 (0.87-1.12) |
| **Admitting specialty**  Medicine*  Surgery**  Family medicine | 1.50 (1.20-1.88)  0.94 (0.73-1.22)  1.00 | 1.16 (0.92-1.46)  1.23 (0.96-1.59)  1.00 | 1.82 (1.37-2.44)  20.8 (15.6-27.8)  1.00 | 1.29 (1.23-1.36)  0.82 (0.75-0.90)  1.00 |
| **Time of handoff**  8:00  16:00  24:00 | 1.00  0.98 (0.73-1.32)  1.01 (0.83-1.24) | 1.00  0.86 (0.64-1.15)  1.21 (0.99-1.48) | 1.00  1.02 (0.81-1.29)  1.04 (0.89-1.22) | 1.00  1.02 (0.90-1.15)  1.05 (0.97-1.14) |
| **Resuscitation bay as initial ED orientation***** | 1.35 (1.11-1.64) | 2.38 (2.00-2.83) | 0.77 (0.65-0.92) | 0.91 (0.84-0.99) |
| **Average number of**  **new ED visits per hour during episode of care per allowed bed** | 0.98 (0.20-4.92) | 0.41 (0.08-2.10) | 0.77 (0.20-2.95) | 1.76 (0.91-3.40) |
| **Average ED occupancy rate during episode of care per allowed bed** | 1.36 (1.11-1.67) | 1.34 (1.10-1.64) | 0.93 (0.78-1.10) | 0.97 (0.89-1.06) |

* Medical specialities include: Internal Medicine and its subspecialties (e.g cardiology), dermatology, medical genetics, neurology, physiatry and psychiatry.

**Surgical specialities include: General surgery and its subspecialties (e.g colorectal), oral surgery, thoracic, cardiac, gynecology, ophthalmology, ENT, orthopedics, plastic and urology

*** For analysis, the ambulatory category and stretcher category were merged as the sample size of the ambulatory category was too small (n=41).
